# “A self-perpetuating malaise in the system:” Understanding scientists’ perspectives on the sex/gender gap in cardiovascular disease research

**DOI:** 10.64898/2026.01.23.26344717

**Authors:** Stephanie E. Coen, Adrian Buttazzoni, Sari Tudiver, Susan P. Phillips, Vivien Runnels, Lorri Puil, Ann Pederson

## Abstract

Longstanding evidence demonstrates that integrating sex-based biological factors and gendered social factors in health research enhances understanding of variation in risk factors, symptoms, progression, diagnoses, and treatments of diseases, and leads to more targeted interventions and improved health outcomes. Sex and gender-based analysis (SGBA) provides a framework for integrating sex/gender and other social constructs and locations throughout the research cycle, addressing the fundamental question: to whom does the evidence apply? Cardiovascular disease (CVD), as the major cause of mortality for men and women globally, and the major cause of premature death in women in Canada, provides a prime example of the gap between evidence that sex/gender matters and the routine uptake of SGBA in practice. This paper addresses some of the transformations needed to overcome this gap by exploring scientists’ perspectives on how sex/gender considerations can be routinely integrated in basic and clinical CVD research. Grounded in key informant interviews with 19 federally-funded CVD scientists in Canada, our thematic analysis generated three themes highlighting how barriers to integrating sex/gender in basic and clinical research practice become embedded in the everyday doing of research: (1) The science of sex/gender gets lost in practice, (2) Institutional and systemic processes maintain the status quo, and (3) Change must centre on accountability and community. While scientists appreciated how SGBA could strengthen research veracity, they identified major obstacles as well as mundane institutional practices and procedures that reinforced the status quo and impeded more varied approaches to research and data collection. Drawing together the ‘bricolage’ of insights, examples, and suggestions provided by the scientists we spoke with, we offer key actions and guidance towards greater inclusivity of sex/gender in the scientific paradigm, and specifically, in CVD research that shapes clinical practice.

## 1. INTRODUCTION

For over four decades, women’s health theorists, advocates, and researchers have demonstrated that integrating sex-based biological factors and gendered social factors in health research can enhance understanding of variation in risk factors, symptoms, progression, diagnoses, and treatments of diseases, and lead to more targeted interventions and improved health outcomes (Doyal, 1995; Greaves, 2015; Harding, 1986; Institute of Medicine (US) Committee on Understanding the Biology of Sex and Gender Differences, 2001; Krieger, 2005; Oakley, 2000). Sex and gender-based analysis (SGBA) provides a conceptual framework for integrating sex/gender and other social constructs and locations throughout the research cycle: in formulating questions; choosing appropriate methods; collecting, generating, and analyzing data; and in discussing the implications of findings, including to whom the evidence applies and to whom it may not apply (Clow et al., 2009; Coen & Banister, 2012). From sex- and gender-sensitive research, we are learning how biological factors associated with gene expression, hormonal function, and reproductive anatomy (sex) are entangled with socially constructed roles, behaviours, and norms that contribute to gendered life experiences, and embodied in physiological and psychosocial conditions, such as autoimmune disorders, Type 2 Diabetes, and chronic pain, often experienced as co-morbidities (Kautzky-Willer et al., 2016; Kinsey et al., 2018; McClements et al., 2025; Osborne & Davis, 2022; Shantz et al., 2024).^1^ However, the integration of sex/gender in health research remains inconsistent, and is subject to confusion and, all too frequently, pushback despite: (i) substantive evidence demonstrating that sex and gender matter in prevalence, presentation, diagnosis, treatment, and outcomes of diseases and conditions; (ii) policy mandates, requirements, and softer incentives on the part of governments, major research funding agencies, and scientific journals to address how and why sex and gender may be relevant to the subject matter; and (iii) accessible SGBA educational resources. In this paper, we explore the perspectives of cardiovascular disease (CVD) researchers to identify barriers and solutions to integrating sex/gender in CVD research, with implications for health research more widely.

### 1.1 Background

At the level of governments, institutes, and agencies supporting health research, there has been more than three decades of formal recognition of the value added by SGBA. For example, The European Commission Directorate-General for Research and Innovation, German Research Foundation, and the US National Institutes of Health (NIH), along with other international partners, have developed innovative initiatives to integrate sex and gender into research in science and engineering (Schiebinger et al., 2025). Canada was an early player in the development of SGBA policies and resources. Health Canada’s Women’s Health Strategy (1999) and Gender-based Analysis Policy (2000), precursors to the current SGBA+ policy (Government of Canada, 2023), articulated SGBA as a systematic, evidence-based analytic tool and framework to be applied to policies, programs, legislation, health surveillance, research and data collection, within their jurisdiction (Des Meules & Stewart, 2003; Government of Canada, 1999, 2023; Health Canada, 2000). Between 1996 and 2012, Centres of Excellence in Women’s Health were federally funded to conduct academic and community-based collaborative research in the social determinants of women’s health, resulting in a body of SGBA-relevant research and resources (Centre of Excellence for Women’s Health, n.d.; Greaves, 2015; Tudiver, 2015). In 2000, the Institute of Gender and Health (IGH) was established as one of 13 institutes of the Canadian Institutes of Health Research (CIHR), Canada’s national health research funder (https://cihr-irsc.gc.ca/e/8673.html). By 2010, all investigators seeking CIHR funding were required to indicate whether sex and/or gender were addressed in their study design, and provide justification for omission (Johnson & Beaudet, 2013; White et al., 2021).

The US National Institutes of Health (NIH), the CIHR, and the European Union (EU) have each developed policies, requirements, educational resources, and training programs to incentivize and support biomedical researchers to implement SGBA. While all cite progress raising awareness about the value of SGBA and integrating sex/gender considerations in research design, they acknowledge the need for accountability and call for cross-sector collaboration to ensure implementation across the research continuum (White et al., 2021, pp. 3045–3046). Measuring such progress is challenging. An analysis of 8,964 abstracts of CIHR Project and Operating grants awarded from 2009-2020 revealed, overall, 91.65% of grants did not have sex or gender mentioned in their proposal abstract, a negligible change from 95% twelve years earlier (Stranges et al., 2023). A prospective, longitudinal evaluation by Haverfield and Tannenbaum (2021) of 39,390 full-text applications across 15 CIHR competitions between 2011-2019 showed a rise from 22% to 83% in applications integrating sex, and from 12% to 33% in those integrating gender, although the terms sex and gender were persistently conflated. Starting in 2018, they also observed applications that integrated sex and gender were more likely to be funded. However, knowing whether sex/gender was addressed, and in what ways, in publications based on this research, would be a more robust indication of sustained progress.

Major health research journals, including the *Canadian Journal of Public Health* (Gahagan, 2016), *The Lancet* (Clark & Horton, 2019; The Lancet, 2025, p. 2), the *Journal of the American Medical Association*, and *BMJ Global Health* (Peters & Norton, 2018; Peters & Woodward, 2023), recommend that prospective authors address sex and gender considerations in their manuscript submissions. The Sex and Gender Equity Research (SAGER) reporting guidelines have played a key role supporting these initiatives internationally (Hawkes & Chang, 2024; Heidari et al., 2011, 2016; Van Epps et al., 2022); adoption of the guidelines by the WHO in 2024 is a further step towards integrating SGBA in international research practices (Heidari et al., 2024). However, a 5^th^ anniversary review of the SAGER guidelines noted growing awareness and adoption but highlighted “critical barriers to systematic implementation of sex-based and gender-based analyses in research and reporting” (Peters et al., 2021, p. 1). These barriers include concerns about time and costs associated with larger sample sizes and additional statistical analyses, editors’ lack of capacity to implement the guidelines, resistance or lack of awareness, and technical challenges. To improve research, the guideline authors called for “structural and systemic changes across the entire research and innovation cycle…involving engagement with universities, professional societies, ethics committees, funders, industry and policy-makers” (ibid, 2).

Past policies that restricted inclusion of women and minoritized populations in clinical trials (Merkatz, 1998; Yakerson, 2019) still influence the limited attention paid to sex/gender in publications across health research sub-specialities. Analysing a sample of 100 Canadian randomized controlled trials of non-sex-specific health subject areas published between January 2013 and July 2014, members of our team demonstrated the near-absence of sex-disaggregated data (4%), of subgroup analyses by sex (6%), and the total absence of discussion about sex/gender differences or similarities, despite relevance of SGBA to the subject areas (Welch et al., 2017). A scientific report published in *Nature* assessing the impact of sex/gender-specific funding and editorial policies on biomedical publications between 2000-2021 in the US, Canada, UK, Japan, China and Korea, showed modest increases in publication output in countries with explicit SGBA policies (US, Canada, UK) over those without. However, biomedical and clinical sciences lagged behind psychology and biological sciences in SGBA-related publications (H. Kim et al., 2024). This disjuncture between policy and published results is not surprising: many granting agencies, including CIHR, do not require grantees to report on whether sex/gender analyses committed to in research proposals were conducted and then included in research findings (Shansky & Murphy, 2021; Stranges et al., 2023).

As the major cause of mortality for men and women globally, and the major cause of premature death in women in Canada, cardiovascular disease (CVD) provides a prime example of the gap between evidence that sex/gender matters and the routine uptake of SGBA in practice. Research shows fundamental sex differences in the anatomy, physiology, and function of the heart from earliest stages of development across the life course (Conlon & Arnold, 2023; P. J. Connelly et al., 2021; Haider et al., 2020; Heart and Stroke, 2023; Pacheco et al., 2022) together with striking gendered inequities including women’s lower participation in and referral to cardiac rehabilitation (Coutinho et al., 2025, 2025). These include variations between men and women in age of disease onset, impact of risk factors, such as hypertension and Type 2 diabetes, prevalence and manifestation of CVD, including coronary artery disease (CAD), cardiomyopathies, heart failure, stroke, arrhythmias and sudden death, and in responses to treatments, including cardiac drugs and medical devices (Bello & Cheng, 2024; Regitz-Zagrosek & Gebhard, 2023; Reue & Wiese, 2022). This knowledge has accrued slowly. For example, women were under-represented in drug trials for heart failure, CAD, and acute coronary syndrome (ACS)/myocardial infarction, while their participation in drug trials for hypertension and atrial fibrillation approached the disease prevalence in women, and exceeded the prevalence in trials for pulmonary arterial hypertension (PAH) (Scott et al., 2018). Despite some improvements, clinical trial participants in major CVD areas do not represent the full spectrum of patients to whom a drug will be prescribed (Lindsey et al., 2021; Pilote & Raparelli, 2018). Further, not only analysis of sex-disaggregated data, but analysis of sex specific data, particularly on CVD conditions affecting pregnant persons, is needed (Geller et al., 2018). Determining whether hypertensive disorders of pregnancy are associated with subsequent heightened risk of CVD, requires collection of sex-specific health data on sex specific risk factors and adequately powered sex-specific analyses (Bello & Cheng, 2024).

In 2021, The Lancet Commission, focusing on women and heart health, identified a state of “stagnation in the overall reduction of cardiovascular disease burden for women in the past decade,” concluding that “cardiovascular disease in women remains understudied, under-recognised, underdiagnosed, and undertreated” (Vogel et al., 2021, p. 2385). A review conducted by the Canadian Women’s Heart Health Alliance at the same time concurred:

> Although the burden of CVD has been improving over time, outcomes for women, particularly over 55, have stagnated. Women are more likely than men to die in the year following an acute myocardial infarction and to experience death, heart failure or stroke within 5 years after acute MI. Women who experience a stroke are at higher risk of mortality, have poorer outcomes, less likely to return home, and also less likely to participate in rehabilitation than men. (Jaffer et al., 2021, p. 1)

The interplay of sex/gender-related factors was striking. For example, women’s lower participation, compared to men, in cardiac rehabilitation is attributed to gender-related factors such as family responsibilities that limit time and access to programs, and/or to lack of information, active encouragement, and lack of referral from health care providers (Coutinho et al., 2025; Khadanga et al., 2021). Indeed, US data suggest a *decline* in women’s awareness about the impacts of CVD, its risk factors and possible symptoms (Nathani et al., 2024), while surveys among Canadian and US primary care providers and cardiologists showed gaps in knowledge and awareness about how to identify and treat heart disease in women (Humphries & Pilote, 2018).

### 1.2 Contextualizing our goals

This paper addresses some of the transformations needed to overcome the stagnation described above by exploring the central question: How can considerations of sex/gender be routinely integrated in CVD basic and clinical research? We build on previous work of our Sex/Gender Methods Group (SGMG) fostering the integration of sex/gender analysis in primary research and research synthesis (https://methods.cochrane.org/equity/sex-and-gender-analysis). For example, we were the first to document the omission of sex/gender in a sample of CVD systematic reviews from the Cochrane Library, demonstrating how omissions in primary studies create limitations in the applicability of synthesised evidence (Doull et al., 2010). We developed SGBA assessment tools for systematic reviews, ‘Briefing notes’ to guide reviewers in addressing sex and gender in areas such as hypertension (Doull et al., 2011, 2014), and published an analysis about the ‘thorny issues’ systematic reviewers encountered when trying to address sex/gender in their work (Runnels et al., 2014). We recognized that addressing SGBA in biomedical health research presented deep-rooted, seemingly intransigent, challenges (Tudiver et al., 2024).

To better understand this intransigence and how it might be addressed, we sought the perspectives and day-to-day experiences of basic and clinical researchers who were principal investigators of federally-funded CVD research grants in Canada. We saw these scientists as valuable key informants who could offer first-hand insight into the complexities involved in addressing sex/gender at the bench or bedside. Our exploration can also be framed in relation to a broader question, pertinent to the history of science: How can relatively new and evolving conceptual frameworks like SGBA, considered marginal to accepted practices, become integrated into, or shift the nature of, a commonly accepted scientific paradigm? Focusing on the perceptions of individual researchers who develop proposals, build research teams, and conduct studies, helps us understand the health research landscape they navigate—a complex web of multiple players, institutions, practices, and policies. Our specific objectives were to identify barriers and facilitators to considering sex/gender at the micro level of basic and clinical everyday research practices and, based on what we learned, to further refine strategies and tools for routine consideration of sex/gender in cardiovascular research.

## 2. METHODS

### 2.1 Key informant recruitment and characteristics

We sought to identify and recruit a cohort of key informants from among CVD researchers working in Canada, diverse by gender and stage of career. This diversity would potentially reveal a wider range of opinions within a relatively small cohort. Given that the Canadian Institutes of Health Research (CIHR) is Canada’s national health research funder, Principal Investigators (PIs) attached to CIHR grants would include active researchers in the Canadian CVD community. We searched publicly available records on the CIHR research database for grants related to CVD research over a period of 5.5 years (January 2012 - June 2017). Since data about the gender of investigators were not available in the CIHR database, gender was initially established by reviewing participants’ professional webpages (e.g., use of pronouns). We stratified the list of grantees by presumed gender and randomized the order of each list. The first 25 from each stratum were invited, via email, to participate in the study, followed by a second round of invitations to the next 20 grantees in each list (90 invitations total). Those who responded with an interest in participating were sent a formal letter that provided background information, an overview of verbal consent procedures, and a copy of the interview questions. By the end of our recruitment period, the final sample of interviewees (*n*=19) comprised 12 active CVD researchers who self-identified as women and 7 as men. Ethics approval was obtained at both the University of British Columbia Children’s & Women’s Hospital Research Ethics Board (H18-00479) and Western University’s Non-Medical Research Ethics Board (111778).

At the start of the interview, participants were asked to confirm their gender to avoid any erroneous inferences based on available online material. Participants included early-, mid-, and senior-career basic and clinical investigators working across a range of CVD fields of study (see Table 1) at universities, hospitals, and research institutions across Canada. To maintain participant anonymity, we categorized their research as either basic or clinical. Basic research included using blood or tissue samples and studies including animals, while clinical research included studies involving human subjects, such as effects of new medications or other interventions (e.g., exercise), and the relationship between CVD and other diseases (e.g., diabetes) or conditions (e.g., pregnancy). When reporting our findings, participant quotes are labelled by the ID code assigned to each participant; within each code, W refers to woman and M to man.

**Table 1.** Key informant characteristics.

| **Participant ID** | **Gender** | **Career Stage** | **Field** |
| --- | --- | --- | --- |
| PW01 | Woman | Senior-career | Clinical |
| PW02 | Woman | Senior-career | Basic science |
| PW03 | Woman | Senior-career | Basic science |
| PW04 | Woman | Senior-career | Basic science |
| PW05 | Woman | Mid-career | Clinical |
| PW06 | Woman | Mid-career | Basic science |
| PW07 | Woman | Senior-career | Clinical |
| PW08 | Woman | Senior-career | Clinical |
| PW09 | Woman | Early-career | Clinical |
| PW10 | Woman | Mid-career | Clinical |
| PW11 | Woman | Senior-career | Clinical |
| PW12 | Woman | Early-career | Clinical |
| PM01 | Man | Mid-career | Clinical |
| PM02 | Man | Mid-career | Clinical |
| PM03 | Man | Senior-career | Clinical |
| PM04 | Man | Senior-career | Basic science |
| PM05 | Man | Senior-career | Basic science |
| PM06 | Man | Mid-career | Basic science |
| PM07 | Man | Mid-career | Basic science and Clinical |
| **Notes:** *Career Stage:* Senior-career ($n=10$), Mid-career ($n=7$), Early-career ($n=2$) \| *\*Field:* Basic science ($n=8$), Clinical ($n=12$) *one participant was both a basic science and clinical researcher \| *Gender:* Woman ($n=12$), Man ($N=n=7$) | | | |

### 2.2 Data Collection

Interview questions were developed by three of the authors and circulated to all members of the team for comments, revisions, and final approval. A semi-structured interview design was chosen as it allows an interviewer to guide the conversations along a series of topics, while also providing flexibility for participants to raise issues important to them (Axinn & Pearce, 2006). Telephone interviews with CVD researchers were conducted by the lead author. Verbal consent was obtained prior to each interview and recorded by the interviewer in an audit log. To ensure commonality in the use of key concepts in our discussions, the interviewer shared the CIHR definitions of sex (e.g., biological attributes) and gender (i.e., socially constructed roles, behaviours, expressions and identities) (Canadian Institutes of Health Research, 2023) with participants and asked if these were consistent with their understandings of the terms; all participants indicated they were. To accommodate the demanding schedules of researchers, many of whom also held clinical duties, we designed the interviews to take between 20 and 30 minutes; they averaged 25 minutes (range: 13-47 minutes). All interviews were audio recorded and transcribed verbatim, removing specific identifying information (e.g., institutional or study name references). Interviews were completed over approximately four weeks in summer 2018.

### 2.3 Data Analysis

Our thematic analysis drew from grounded theory principles in our inductive approach to identifying patterns in the data (Glaser, 2002; Thomas and James, 2006). A systematic iterative coding process was undertaken by three authors (SEC, AB, AP). First, two authors (SEC, AB) independently completed line-by-line coding of all the transcripts, identifying both commonly occurring ideas and sentiments, as well as significant departures and tensions, to build a set of codes (Auerbach & Silverstein, 2003). Next, the coding team explored possible interpretations and relationships amongst the codes, leading to agreement on nine robust codes that were organized into three higher-order themes. We strived for ‘multivocality’ (Tracy, 2010), paying close attention to the nuances, contradictions, and unique expressions within a single interview, as well as to possible patterns and disjuncture between and among the varied voices represented in our findings (Roger et al., 2018). To emphasize and honour the rich detail and analysis that was shared, we adopted an *in vivo* approach, using participants’ own words to elaborate themes and to name codes (Manning, 2017). Our research methods may be characterized as a form of ‘bricolage’, carefully mining the diverse answers we were given for the levels of analysis, perspectives, and insights they offer, then stitching them together into a patchwork whole (Kaufert, 2000). Finally, our thematic framework was presented to the entire research team, who engaged in what Smith and McGannon (2018) outline as the practice of review by ‘critical friends’ to interrogate and clarify our interpretations. An overview of our coding framework and theme construction is provided in Figure 1.

**Figure 1.**
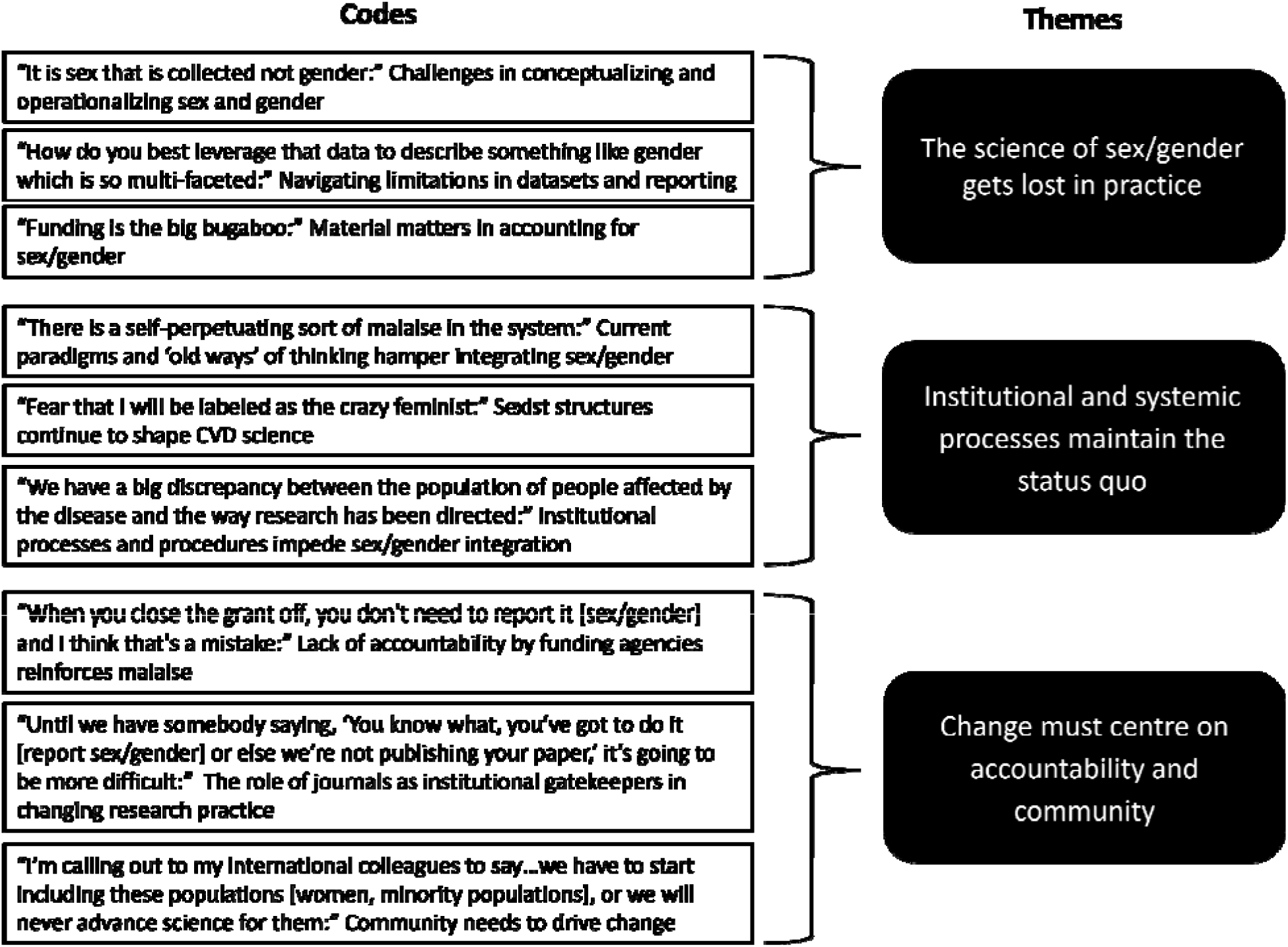
Overview of coding framework and theme construction.

## 3. RESULTS

The three themes generated in our analysis highlight how barriers to integrating sex/gender in basic and clinical research practice become embedded in the everyday doing of research. The first theme, **‘The science of sex/gender gets lost in practice,’** captures how scientists grapple with fundamental conceptual, methodological, and logistical challenges that complicate the routine integration of sex/gender. The second theme, **‘Institutional and systemic processes maintain the status quo,’** reveals how material and socio-cultural structures entrench long-standing ways of researching CVD. Finally, **‘Change must centre on accountability and community,’** sheds light into some of the gaps researchers perceive between policy mandates and implementation of these requirements, and highlights strategies for change.

### 3.1 Theme 1: The science of sex/gender gets lost in practice

This theme reflects the discordance between, on the one hand, scientists’ knowledge that sex and gender matter in CVD prevalence, presentation, treatment, and outcomes and, on the other hand, their difficulties in conceptualising and operationalising sex and gender in specific research practices. Only one participant, a mid-career basic scientist, said he encountered no barriers to considering sex/gender because he located those influences beyond his immediate field of study: “I think it’s absolutely incumbent upon us to study those differences, so I do believe that that has to be done. But that’s further downstream than what I’m doing” (PM06).

More commonly, participants described a range of obstacles to considering sex and/or gender in their research. Several noted the “tendency to wrongly use sex and gender interchangeably” (PW11), not only in relation to CVD research, but across the health sciences. This conflation of sex and gender is widely noted in the literature and leads to confusion over what exactly is being characterised, analysed, or measured (Kaufman et al., 2023; Keener, 2015). The lack of clarity and consistency in terminology was further compounded for CVD researchers working in the bilingual context of Canada where sex/gender and *sexe/genre* carry different nuances and present translation challenges for already complex concepts. For example, even Canada’s CIHR Institute of Gender and Health is translated into French as Institut de la santé des femmes et des hommes (literally, the Institute of Men’s and Women’s Health). One scientist addressed how these difficulties in translation could further complicate and even increase resistance to the uptake of sex/gender in scientific work:

> …I think that the vast majority of people in the world do not understand the difference between sex and gender, and I’m in Quebec—and I think a lot of Canada starts to get it [the difference]—I think in Quebec it’s really hard and that’s because, you know [hesitate], sex versus gender, just as terminology in Quebec is different. Right? So, so not only that the words aren’t the same kinds of words, and so trying to get across the nuance between these two is just, is really complex. So anyways, I think there is a huge amount of confusion, a huge amount of resistance to thinking about these in those ways. (PM02)

The same basic scientist also described trying to explain the concept of gender to colleagues:

> …you know, I can sit there and talk about how gender is this sort of psycho-social cultural phenomenon, and that’s very easy. I can almost be a little bit glib about that. People might say, ‘okay, what does that mean?’ Then you’re like, yeah, well it’s multilayered, multi-tiered, and there’s so many angles to it—and then a researcher goes to me, quite legitimately, and says ‘how do I measure it?’ And I’m like, ‘I wish I had a good answer for you’ [laughing]. (PM02)

A mid-career clinical scientist probed the very meaning of the constructs, questioning whether the terms served as proxies for processes that are not clearly identified, nor measured.

> I often wonder about people using sex or gender as a proxy measure for what they’re actually really interested in. So, we might say gender is what influences participation in physical activity, but maybe it’s not so much the person’s gender as it is the social roles that are associated with that [their gender]—that are typically more associated with one gender or another. (PW05)

These comments suggest, for some, use of the terms sex and gender may actually obscure understanding of the underlying processes. They also raise the question as to *which* aspects of gender matter to the outcome of interest—for example, is it gender identity at an individual level or gender roles at a structural level, or the interplay of these? We return to this concern in the discussion section of the article.

The investigators described the ways clinical research is embedded in pre-determined practices, structures, and datasets that constrained more nimble, nuanced considerations of sex and gender. This resulted in under-representation of women in specific areas of CVD clinical trial research and also extended to minoritized populations. A senior clinical scientist reflected on how under-recruitment in clinical trials can affect clinical treatment decisions and health outcomes:

> Even when you look at the clinical trials that went down in the past they have still been dominated by men in terms of enrolment. White men in terms of enrolment. So, this is a real problem. Since the time when it was starting to be mandated that you have to address sex and gender in your applications for Heart and Stroke (Foundation) and CIHR, for example, still two thirds of the clinical trials in cardiovascular disease are in men. […] so, we don’t really know if some of the therapies that are guideline recommendations, actually work in the real world, especially in women. (PW08)

Reliance on hospital-based medical records to identify potential human research subjects could also mean that study populations are an artefact of sex/gender biases in treatment referrals. A mid-career basic scientist considered how these dynamics may shape what is observed and studied in the clinical context:

> Sex as a variable is definitely more commonplace in sort of the biomedical basic science research. But indirectly, I think that gender impacts in the clinical care eventually trickle down to us in terms of what diseases are actually being seen in the clinic, and then how do we understand that disease and how it may be different. (PW06)

This researcher recognized a chain of effects across the research and clinical care systems influencing who is studied and therefore what is learned about sex and gender similarities and differences. In addition, recruitment of potential research subjects through hospital-based medical records restricted clinical samples to binary sex-based categories. An early-career clinical scientist reflected how this might limit the potential for more complex considerations of sex and gender:

> We don’t really assess gender very thoroughly from a clinical point of view. […] Any retrospective trials, or even prospective trials, when we’re working with our clinical programs, the information that would be sent to us is purely male versus female. And that is how we would then capture male versus female in publication. And also perform analyses from a purely biological point of view. (PW09)

Investigators raised fundamental questions about the current capacity of methodologies and specific tools to empirically account for sex/gender, from cellular to societal levels. The mid-career basic scientist quoted above considered whether cells remain sexed once they have been cultured in petri dish environments over time:

> When you get [cells] from like a human donor you know that human donor was male or female. But sometimes you know people have been using cell lines that have been ‘in culture’. I don’t know if you’ve ever heard of like HeLa cells, like Henrietta Lacks—there’s been one cell type that’s been ‘cultured’ from around the world for decades and decades and decades. Obviously, she was a woman. Are the HeLa cells still female? Obviously, nobody knows the answer to the questions. Nobody knows what makes a cell in a dish male or female other than obviously the number of Y chromosomes or whatever. (PW06)

Others queried whether certain tools and constructs were fit for purpose due to changing gender roles and norms of masculinity and femininity. A mid-career clinical scientist challenged the relevance of the BEM Sex Role Inventory, developed in the 1970s (Bem, 1974), “It kind of blows my mind, how do we best measure [gender]… and develop clinical interventions and things that will allow you to improve the health of the nation with something [gender] so quickly shifting?” (PM02)

Alongside conceptual and methodological challenges, for many scientists “funding is the big bugaboo” (PM05), creating practical and logistical barriers to adequately addressing sex/gender in their research. Discrepancies between what sex/gender funding policies demanded versus what was actually funded often impeded researchers’ capacity to integrate sex/gender in their work—that is, granting agencies “have nice principles, but they don’t facilitate the research” (PW02). A senior basic scientist explained:

> I put my proposal in and I say I need $1.2 million over 5 years and they give me $900,000. I’m really grateful for it. But, $900,000 is not $1.2 million, and that $300,000 is going to be felt in the study design, and I suspect that for many people one of the first things that will go is an analysis [of] sex and gender considerations. (PM05)

While many scientists held the view that accounting for sex, particularly, in animal studies, could double study costs, a mid-career basic scientist disagreed:

> Everyone actually thinks that, ‘Well, if I’m going to have to do everything in both males and females, you’re going to have to double everyone’s budget.’ And that’s actually not true because, for example, like all of my sample analyses don’t cost like exactly one dollar per analysis and then therefore I have to do two analyses. Like, there are lots of aggregated costs. (PW06)

Regardless of whether actual costs ‘doubled’ or not, the mismatch between funding requested and funding received created a disconnect between sex/gender policy priorities and on-the-ground research capacity, signaling a mixed message to the research community.

### 3.2 Theme 2: Institutional and systemic processes maintain the status quo

This theme identified how underlying institutional and systemic processes limit attention to sex/gender in CVD research and clinical practice. One senior basic scientist pointed to “a self-perpetuating sort of malaise in the system” (PW04) as a fundamental impediment to transforming the culture around sex/gender in CVD research. Part of this malaise, she explained, were undercurrents or unstated assumptions in relation to sex/gender that “because something is not known it’s interpreted as something that is not worth knowing” (PW04). This could create a “kind of a catch-22,” where scientists felt constrained in exploring sex and gender-related questions because “nothing is known and therefore we don’t have a strong enough basis to ask [sex/gender] questions.” She also shared her “fear that I will be labeled as the crazy feminist,” noting “it’s easier to be silenced if you are in a small minority” (PW04).

Similarly, a senior clinical scientist described being the only woman amongst a group of men researchers discussing sex/gender inclusion policies; the majority response was, “until we know it’s important, then we shouldn’t require researchers to include it in the studies” (PW07). Another senior clinical scientist described her struggles to challenge prevailing orthodoxies in clinical teaching settings:

> It [translating sex/gender implications into clinical settings] is a challenge and it’s an interesting thing because cardiology is a male-dominated specialty that’s slowly changing. But certainly, when I started in the field, it was a very male-dominated specialty. So, you get on your soap box, and you try to explain that women will present differently and you try to educate the trainees that women present differently but still have these risks and still have these underlying challenges, and I think it’s better than it was, but it’s still a problem. (PW08)

Stroke interventions offered insights into the value of SGBA. A mid-career clinical scientist detailed how the long-standing ‘time-bound’ paradigm used to identify and treat acute stroke, while effective for many patients, has also contributed to poorer outcomes and higher mortality in women than in men. As he explained, the ‘time-bound’ approach of using a window of 4-6 hours from the start of stroke symptoms as the sole criterion for thrombolytic intervention resulted in missed opportunities, particularly in women whose arrival at hospital was often delayed relative to men’s. As this scientist explained, “men are not witnessing women’s strokes because they are not around to witness it. And what happens then is our own stroke treatment paradigm is time-bound, I guess, you know, women are under-treated” (PM01). He went on to describe the value of acute stroke imaging and a measuring-based treatment paradigm:

> We were actually wanting to move our field away from time-based paradigms to tissue or measuring-based paradigms, which means that somebody comes into the hospital - you don’t actually deny people treatment based on witnessed stroke or known or unknown onset time, you scan them, as a sign of time entry. (PM01)

Other researchers addressed how inequities in access to treatment also perpetuated inequities in who was included in certain types of clinical research. According to an early career clinical research scientist, because fewer women were referred for CVD-related rehabilitation treatment, fewer women were recruited into clinical trials and other studies that commonly drew participants from these treatment programs. She mapped out the circuitous nature of this problem as reflected in her clinical research setting:

> The majority of the participants that enter and complete [the program] are men. By the nature of the cardiac rehabilitation program, we recruit most of our participants from our existing clinical cardiac rehab program. So, we do not stratify by sex or gender. And we do not aim to recruit the same number of women or men as a result of the few women that enter and complete our program. So, when we publish our data, we clearly publish the number of females and the number of males in our trials, and we routinely will comment in the discussion of how fewer women or females than men are included in these trials. But, that’s really a reflection of our clinical programs in general: they’re not meeting the needs of women. (PW09)

In the basic sciences, researchers working with human tissue samples encountered various institutional impediments to addressing sex. A senior clinical scientist (PM03) explained that the sex of human tissue donors could be unavailable to researchers due to ethical regulations obligating anonymity in human samples: “We have to assure the anonymity of the donors and have no information regarding age or sex.” He went on to lament his experience that “unfortunately, we cannot say ‘Ok, could you please give me 5 female 5 male donors’, because it is a bad system in that way.” This highlighted the need to reconcile institutional privacy policies and other necessary ethical practices with the value accrued through sex-specific data.

These comments from basic and clinical scientists called attention to mutually reinforcing systems and procedures that limited and, at times undermined, opportunities to consider sex/gender in the conduct of CVD research and in clinical practice. Within these settings, researchers described obstacles to addressing potential sex/gender-related differences in their research questions and recruiting representation from diverse populations experiencing cardiovascular diseases, leading to gaps in the research evidence informing clinical decisions and treatment guidelines. As one senior clinical scientist commented, “We have a big discrepancy between the population of people affected by the disease and the way research has been directed” (PW08). Another researcher drew the link between sex/gender and other intersectional factors and processes:

> And, where we land on the sex/gender issue, we can take very similar things and translate it to people with lower socioeconomic status and people who have immigrated to the country, Indigenous people, because none of this has been represented in clinical trials, right? (PW08)

All the scientists interviewed described seemingly mundane, entrenched, practical barriers to addressing sex/gender. We turn now to consider what the scientists saw as emerging opportunities and necessary conditions to change the status quo.

### 3.3 Theme 3: Change must centre on accountability and community

When asked how to improve the integration of sex/gender in CVD research, investigators spoke about two complementary approaches. First, they identified the need for policies with clear measures of accountability, and for these policies and measures to be harmonised across realms with high academic currency, specifically, research funding agencies and academic journals. These institutions inform new directions in basic and clinical research and clinical practice, and impact professional career advancement. Second, many investigators pointed to ways that they and others within the CVD research community could take a more active role in driving change.

Several interviewees captured tensions inherent in integrating sex/gender in CVD research: while one noted funding agency policies mandating inclusion of sex/gender in grant proposals are “actually making people think about these issues” (PM01), another raised concerns that “making it a broad stroke for everybody to do sex and gender might also be disingenuous,” running the risk of positioning sex/gender as a “flavour of the month” (PW03). Still another labelled the requirement to include sex and gender considerations in proposals as tokenism, “because it will tick a box in the review form, and that can be as detrimental as not covering it at all to begin with” (PM05). A senior clinical scientist emphasized the lack of accountability within agency policies, noting that “in the end, when you close the grant off, you don’t need to report it [sex/gender] and I think that’s a mistake. That’s a lost opportunity” (PW01).

One senior basic scientist, whose research dealt specifically with sex differences, advocated for a phased approach that first studies a phenomenon in one sex and then considers sex differences, noting “there’s no point of wasting all that money, all these animals, all the time of your people in your lab to do this right at the beginning of the project” (PW02). Despite this caveat specific to her own laboratory work, she acknowledged the overall benefits of current policies, and echoed the sentiments of most other scientists we spoke with when she said, “We may be exaggerating right now on the emphasis of everything, but I think it’s for the best. Eventually we’ll find the right medium” (PW02). Regardless of granting agency policies, journals could ultimately determine how the science gets done, and were singled out for the role they play in fostering cultural and behavioural shifts in the research community. Participants offered a variety of editorial policy suggestions that journals could adopt, such as requiring authors to “at least, at minimum, report one sentence” (PM04) on the sex/gender characteristics of samples. Others suggested more complex, incentive-based approaches:

> The other thing would be to actually have incentives, and by incentives I mean deterrents to not doing it [reporting sex/gender] […] as researchers that really is our currency—what we publish—until we have somebody saying, ‘You know what, you’ve got to do it [report sex/gender] or else we’re not publishing your paper,’ it’s going to be more difficult to justify and to get into it. So, it’s not an incentive so much as disincentive not to [report on sex and gender]. That would be a huge one. And to actually follow through, because of course they all say it, every single one of the journals say it in our guidelines, but no one actually—there’s no consequence to not following it. Same with the granting agencies. (PW06)

This follow-through in editorial policy related to sex/gender would also need to be applied in the peer review process. A senior clinical scientist recounted how, having integrated sex/gender analysis in a paper submitted for publication, he received peer review recommendations that conflicted with sex/gender editorial policies: “We got back comments from reviewers that said ‘why do you bother with male and female? Remove one” (PM03). A mid-career clinical scientist also spoke to the importance of integrating funding and publishing policies concerning sex/gender to ensure accountability and transparency. He emphasized the need for funding agencies to identify and support those who do include such analyses in their reporting: “You should direct the funding agency to where gender and age-based analysis was reported… People say in the grants application, they’re going to do all this wonderful crap, so they can win and they never do it and then never get caught at the end” (PM01). Researchers consistently suggested that amending and enforcing publication requirements in scientific journals would be a highly effective strategy for promoting the inclusion of sex/gender in future CVD research.

Participants agreed it was incumbent upon individuals within the CVD research community to be agents of change in addressing sex/gender. This could take many forms, from speaking up in informal settings about the value of sex/gender analyses in their own work to advocating for considerations of SGBA in peer-review processes and for inclusion of women and minoritized populations in research:

> I’m appealing or I’m calling out to my international colleagues to say…we have to start including these populations [women, minoritized populations], or we will never advance science for them. I think one way we do that is peer-reviewed research so that regulatory agencies will, I think, over time, will become more inclusive. But it’ll take a drive from us, from the community of scientists. We have to start this. (PW01)

While most researchers struggled with the conceptual complexities and methods of analyzing sex/gender, many gave mixed reviews of how well existing educational tools equipped them for doing the work. Rather than additional resource training, some participants preferred being part of research teams that included those with expertise in SGBA: “My strategy is going to be to collaborate with someone who’s an expert in sex-dependent cardiovascular disease because I think that’s going to be more credible and more effective than me doing a workshop and trying to write a proposal that’s based on that” (PM07). Several researchers provided examples of current practices that may help integrate sex/gender considerations into future research and reporting. A senior basic scientist consistently reported on the sex of research samples:

> …looking through the literature, you know, and getting to this point where a lot of work that we’ve done in the ‘80s and ‘70s, may actually be completely useless in the long run, to the people who will look at this in 50 years from now, because nobody reports ‘what about the sex of your assay?’ So, what we’ve done now, sort of religiously do at least, one thing in all of our publications—we don’t get into really distinguishing the sex in the context of our conclusions but—we say ‘this was done with the cells that were female, and this was done with the cells that were male.’ And so, hopefully in the future somebody who’s looking at that [our findings] can say ‘okay, well, that’s useful information for my studies on sex difference and gender difference’. (PM04)

The scientist saw consistent reporting on the sex of cell lines as a necessary, if minimal, requirement to ensure the replicability and applicability of their team’s scientific work.

## 4. DISCUSSION

### 4.1 Overcoming malaise

The scientists interviewed appreciated how SGBA could strengthen research veracity. They also offered insights into major obstacles, contradictions, and disconnects encountered in trying to address sex/gender in CVD research. Many pointed to everyday, seemingly mundane, settings and routine institutional practices and procedures that reinforced the status quo and impeded more varied approaches to research and data collection. The comment from a senior investigator about a “self-perpetuating sort of malaise in the system” pointedly captured the entrenched nature of these barriers: long-standing paradigms, frameworks, and prevailing orthodoxies that excluded or negated sex/gender in the formulation of research questions and in the design of methodologies; strategies for recruiting research subjects that did not consistently reflect the biologic and social diversity of those experiencing the cardiovascular conditions being investigated; published research that sometimes failed to gather and often failed to report on the sex of cells, tissues, or human participants. Reporting on how gender matters was even more sporadic with some confusion about indicators to use, particularly related to the multi-layered nature of gender (i.e., from individual gender identity to wider gendered social structures like gender roles). These ambiguities and omissions are part of interlocking structures that often do not require, support, or reward inclusive and intersectional research.

Elements of this malaise persist despite policies from governments, granting agencies, and journals mandating the application of SGBA to health research and many robust SGBA implementation initiatives. Some participants revealed an unresolved tension in how they perceived SGBA in relation to scientific inquiry. On the one hand, they supported policies mandating SGBA be addressed in research grant proposals and scientific publications; on the other, some of the same investigators advocated for an approach to ‘go where the science leads,’ hesitant to apply SGBA ‘in a rote way.’ In these comments, SGBA was positioned as a potential obstacle to the pursuit of science and to researcher autonomy, rather than an analytic framework integral to the scientific method and a means to strengthen the potential applicability of research. Investigators saw SGBA likely to be among the first areas scaled back by researchers when granting agencies provided fewer funds than requested.

Malaise was also reflected in resistance to change. Participants who wanted to integrate sex/gender in their CVD research or clinical training described settings in which they felt silenced and encountered pushback. Sometimes subtle, sometimes more overt, sexism embedded in attitudes and behaviours of colleagues also reinforced fears, particularly for women, of not being taken seriously within their profession.

It is hardly surprising that attempts to include sex/gender in research encounter resistance. As described in studies of organizational culture and change, implementation science, and gender mainstreaming, governments, health organizations, and other complex institutions are often characterized by silos of entrenchment and inertia (Graham, 2021; Lombardo & Mergaert, 2013; Moser & Moser, 2005). These dynamics are reflected, for example, in a lack of harmonization among surveillance and other datasets, such as those gathered by provincial and federal health systems (Hannay, 2025) and in poor coordination among departments responsible for implementing major policy initiatives to improve population health and well-being. Efforts to realign, harmonize, and transform long-standing paradigms, practices and procedures, and to change attitudes, are met with passive or active resistance at various levels within public and private sector organizations for many reasons. Key players may push back, fearing heavier workloads, competing priorities, or lack of resources; some individuals may be reluctant to engage in a new field of study, and/or fear loss of personal influence and control. Overcoming pushback depends on many factors: organizational, structural, and environmental support for change, including finances, time, relevant knowledge, effective leadership that can motivate “buy in”, the elusive notion of ‘organizational readiness’ and the broader contextual milieu (Bonawitz et al., 2020; Gadsden et al., 2024; Nilsen & Bernhardsson, 2019). However, malaise can be overcome readily when it is seen as imperative to do so; in the face of the COVID-19 pandemic, individuals, governments, pharmaceutical and other industries mobilized resources to reshape common practices and responded creatively and collaboratively to rapidly-evolving situations.

SGBA is a powerful lens through which to frame research questions and re-view policies and systems to ensure best scientific and clinical practices (Rich-Edwards et al., 2018; Ritz et al., 2014; Tannenbaum et al., 2019). Recent initiatives to develop guidelines for CVD research are heeding the call to action (Usselman et al., 2024). Drawing together the ‘bricolage’ of insights, examples and suggestions provided by the CVD scientists we spoke with, we offer the following guidance towards greater inclusivity of sex/gender in the scientific paradigm, and specifically, in the CVD research that shapes clinical practice (Table 2).

**Table 2.**
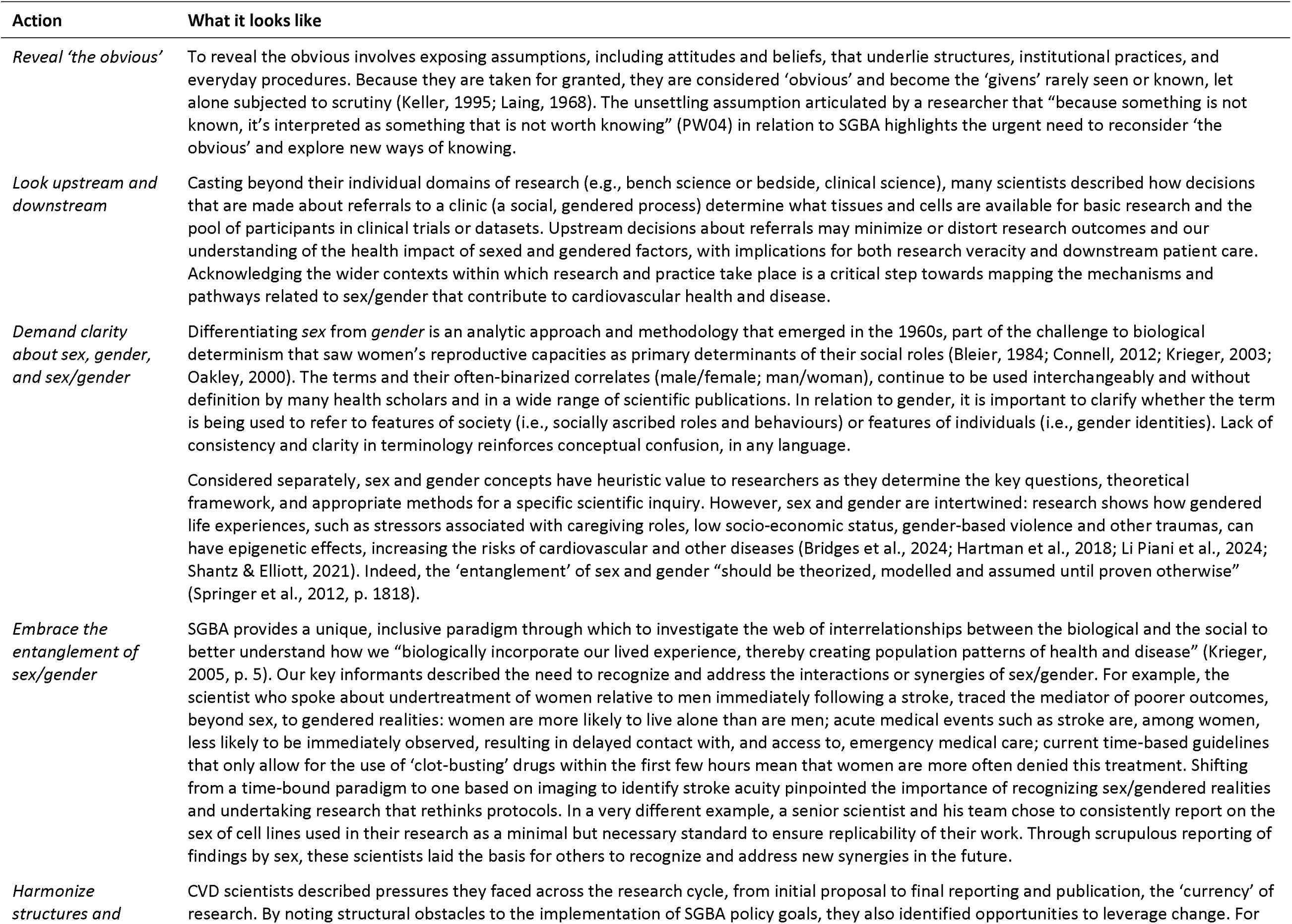
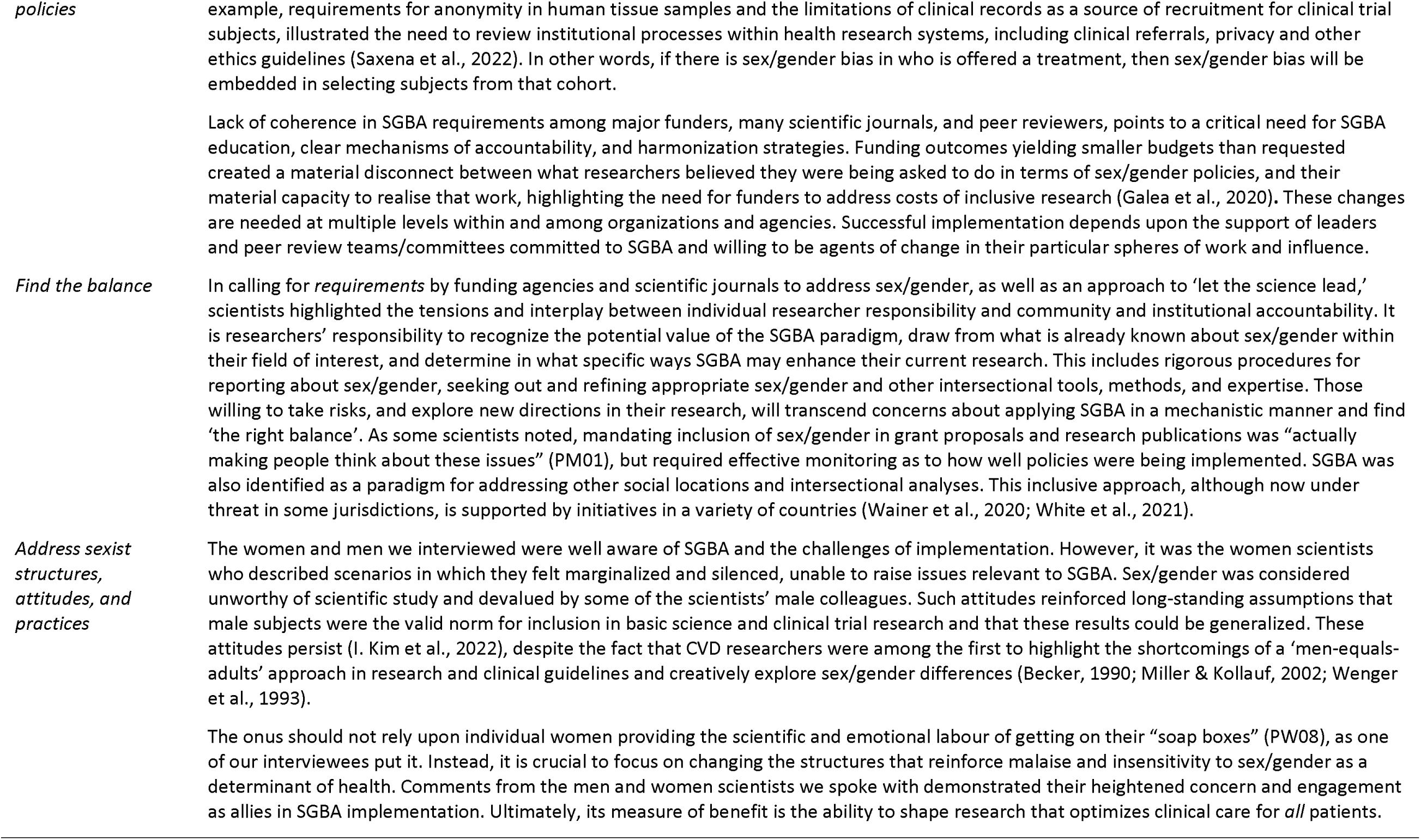
Actions towards greater inclusivity of sex/gender in the CVD research paradigm.

### 4.2 Limitations

This paper has several limitations. First, our findings are based on semi-structured interviews with CVD researchers all working in basic science and/or clinical research settings. As a result, our conclusions are most relevant to CVD basic science and clinical contexts, rather than other areas of CVD research (e.g., health services). Second, given our recruitment process, participants may have self-selected due to a pre-existing interest in sex/gender. To explore issues of malaise and resistance in greater depth, future research might purposefully engage with researchers less inclined to consider sex/gender in their work. That said, those who chose to participate were more likely to ‘see’ the sex/gender landscape within CVD research. Third, while we address sexism as an obstacle to implementation of SGBA in CVD research in the discussion, our methodology was not designed to determine whether there were significant differences between women and men in how they perceived the integration of sex/gender in CVD research and practices. Fourth, this article has largely considered sex and gender in binary terms (men/women; male/female) and assumed cisgender populations. This was due to the overwhelming focus on sex within CVD basic science and clinical research and common assumptions about gender as a binary within the institutions supporting that work. Finally, interviews were conducted in 2018. Our review of recent CVD literature suggests that there has been no meaningful change in funding and that despite initiatives to address sex/gender in journal publication policies, recognition of the merit of sex/gender inclusive research has stagnated or diminished in relation to CVD. As a result, the time lag between data collection and presentation has evolved into a strength in demonstrating a longer-term perspective.

## 5. CONCLUSIONS

The unstated assumption about SGBA, articulated by a senior scientist, that “because something is not known it’s interpreted as something that is not worth knowing” (PW04) is embedded in many of the practices of health systems and in the attitudes of individuals that sustain them. It is an assumption antithetical to sound science and to exploring new directions in research. As a ‘fault line’ (Papanek, 1985) that cuts across all social locations, sex/gender has the potential to enhance and deepen our understanding of issues, to explore the entanglements of the biological and the social by asking questions about applicability and context. Failure to do so results in lost opportunities to advance scientific inquiry and improve the quality and sensitivity of research and clinical interventions. The current political backlash in the U.S. is attempting to erase SGBA-related questions and concepts, and the inclusivity they represent (E. Connelly, 2025; Tanne, 2025). More than not worth knowing, the freedom to ask and explore answers to these questions are under attack. It is prudent and necessary to strategize about how to effectively confront refusals to recognize the significance of sex/gender, and the underlying rejection of science itself.

## Data Availability

Data produced in this study are not available as participants did not consent to data sharing.

## Acknowledgements

We thank the CVD scientists who so generously shared their time and insights with us. This project was part of the overall mission and work of the Sex/Gender Methods Group (https://methods.cochrane.org/equity/sex-and-gender-analysis) and benefited from the support and feedback of all Group members, especially Vivian Welch. Additional thanks to Dr. Jason Gilliland who supported and enabled SEC’s leadership on this project during her postdoctoral fellowship in the Human Environments Analysis Laboratory (Western University, Canada).

## Authors’ contribution statement (CRediT)

**SEC:** conceptualization, funding acquisition, data curation, formal analysis, investigation, methodology, project administration, supervision, writing – original draft preparation; **AB:** data curation, formal analysis, investigation, writing – original draft preparation; **ST:** conceptualization, funding acquisition, formal analysis, writing – original draft preparation; **SPP:** conceptualization, formal analysis, writing – original draft preparation; **VR:** conceptualization, funding acquisition, writing – review & editing **LP**: conceptualization, funding acquisition, writing – review & editing; **AP:** conceptualization, funding acquisition, formal analysis, investigation, methodology, project administration, supervision, writing – original draft preparation.

## Footnotes

1 We use ‘sex/gender’ throughout this manuscript to acknowledge the interactive influences of sex and gender, while referring to ‘sex’ and ‘gender’ specifically where relevant. We also note that while we are focusing on sex-and gender-based analysis (SGBA) here, sexed/gendered processes invariably intersect with other structures of power and marginalization (Government of Canada, 2023).

